# Association of Initial Clinical Characteristics with the Need for the Intensive Care Unit and Hospitalization in Patients Presenting to the Emergency Department with Acute Symptomatic COVID-19

**DOI:** 10.1101/2021.09.19.21263800

**Authors:** Benjamin Zollinger, Sophia Newton, Jincong Q. Freeman, Seamus Moran, Nataly Montano Vargas, Yan Ma, Andrew C. Meltzer

## Abstract

**Objective:** To evaluate the association of initial clinical symptoms with need for hospitalization, intensive care, or death in ED patients within 30 days after presenting with acute symptomatic COVID-19.

**Methods:** This study is a retrospective case-series of patients presenting to a single ED with acute symptomatic COVID-19 from March 7–August 9, 2020. Symptomatic patients with laboratory-confirmed SARS-CoV-2 infection were eligible for this study. Patients who tested positive for COVID-19 due to screening tests but had no reasonably associated symptoms were excluded. Participants were analyzed by three categories representative of clinical severity: intensive care unit (ICU) care/death, general ward admission, and ED discharge/convalescence at home. Outcomes were ascertained 30 days after initial presentation to account for escalation in severity after the ED visit. We conducted univariate and multivariable logistic regression analyses to report odds ratios (OR) with 95% confidence intervals (CI) between hospital or ICU care/death versus convalescence at home and between ICU care/death versus general ward admission.

**Results:** In total, 994 patients were included in the study, of which, 551 (55.4%) patients convalesced at home, 314 (31.6%) patients required general ward admission, and 129 (13.0%) required ICU care or died. In the multivariable models, ED patients requiring hospital admission were more likely to be aged ≥ 65 years (adjusted OR [aOR] 7.4, 95% CI: 5.0, 10.8), Black/African American (aOR 3.0, 95% CI: 1.6, 5.8) or Asian/American Indian/Alaska Native/Other (aOR 2.2, 95% CI: 1.1, 4.3), and experience dyspnea (aOR 2.7, 95% CI: 2.0, 3.7) or diarrhea (aOR 1.6, 95% CI: 1.1, 2.2). However, they were less likely to experience sore throat (aOR 0.4, 95% CI: 0.2, 0.6), myalgia (aOR 0.5, 95% CI: 0.4, 0.7), headache (aOR 0.5, 95% CI: 0.4, 0.8), or olfactory/taste disturbance (aOR 0.5, 95% CI: 0.3, 0.8). ED patients who required ICU care or died were more likely to experience altered mental status (aOR 3.8, 95% CI: 2.1, 6.6), but were less likely to report history of fever (0.5, 95% CI: 0.3, 0.8).

**Conclusions:** COVID-19 presents with a multitude of clinical symptoms and an understanding of the association of symptoms with clinical severity may be useful for predicting ultimate patient outcomes.

## 1 INTRODUCTION

The first case of COVID-19 was reported in the US in January 2020. As of June 4^th^, 2021, there have been 33.1 million cases of COVID-19 infection with over 590,000 deaths in the US. [1] The clinical course ranges widely in severity from asymptomatic to life-threatening. The disease can manifest with a heterogenous constellation of symptoms over its course, and symptomatic patients may present with one or more symptoms. [2-5] Understanding which symptoms at initial presentation are associated with a severe outcome may assist targeting of early intervention and resources. In addition, a systematic understanding of disease course may inform prognosis and resource allocation which is especially important during patient surges. The objective of this study was to analyze the association of initial clinical symptoms present in the ED with the clinical severity of COVID-19.

## 2 METHODS

### 2.1 Study Design and Setting

This was a single-center retrospective case series of patients at an academic urban hospital ED in Washington, DC with approximately 80,000 annual visits. The protocol was approved by the Institutional Review Board at George Washington University with a waiver of consent on March 30, 2020.

### 2.2 Study Population

Patients with a positive nasopharyngeal swab for SARS-CoV-2 by polymerase chain reaction (PCR) on an index ED visit or within 14 days prior to the ED visit were included in this analysis. Patients were excluded if there was a high degree of certainty that the visit was not related to a viral infection such as presentations due to trauma, alcohol or drug intoxication, poisoning, suicidality, suspected rape or other domestic violence, involuntary commitment, or other isolated chief complaint clearly not related to COVID-19. Furthermore, patients who incidentally tested positive due to institutional routine testing but who displayed no COVID-19 or other viral symptoms were also excluded. No asymptomatic patients were included in this analysis. Clinical care decisions to discharge from the ED, admit to the general medical service, or admit to ICU were made according to individual physicians using professional judgement, and are typically based on a patient’s clinical stability, oxygen or ventilator requirements, or nursing supervision requirements.

### 2.3 Data Collection and Processing

Patients were identified via electronic health record (EHR) reviews at least 30 days after their index visit from March 7 until August 9, 2020. The index ED visit was designated as the first presentation of the patient to the George Washington Hospital ED where they tested positive for COVID-19. For all subjects identified, data abstraction was performed by querying the EHR (Cerner™) at least 30 days after their index visit or after the patient was discharged from the hospital, whichever was longer. Chart review was performed according to guidelines by Gilbert et al. [6] The case report forms were created by Indiana University for use in a nation-wide multi-center patient registry. All data was entered into a secure REDCap database. Data abstractors were trained specifically for EHR queries for this study, and 10% of patient charts reviewed were cross-checked by a second abstractor for accuracy.

### 2.4 Statistical Analysis

Data was initially stratified to identify significant associations per three categories of primary outcome: discharge from the ED/convalescence at home, admit to the general ward admission, admit to ICU-level care/death. Patients who were initially discharged but then returned to the ED and needed admission within 30 days were classified in the hospital admission category. Patients who were first admitted to the general wards but ultimately were transferred to ICU were included in the more severe category. Any death within 30 days was categorized in the most severe category.

Clinical symptoms recorded include sore throat, dry or wet cough, chest pain, respiratory distress/failure, history of fever, fatigue/malaise, myalgia, headache, altered mental status (AMS), nausea, diarrhea, olfactory taste disturbance, and syncope. Demographic characteristics included age, sex, race, and ethnicity. Summary statistics were used to describe the demographics and initial clinical symptoms of patients per disposition. Frequencies and proportions were tabulated for categorical variables, with p-values calculated using Chi-square or Fisher’s exact tests. Means and standard deviations were reported for continuous data, with p-values calculated using analysis of variance (ANOVA).

Prior to regression analysis, variables were selected based on clinical knowledge and with a *p*-value <0.1 from bivariate analyses. We conducted univariate and multivariable logistic regression analyses to report odds ratios (OR) and adjusted odds ratios (aOR) with 95% confidence intervals (CI). We ran two separate logistic regression models: hospital admission (inclusive of ICU level patients) versus convalescence at home and between ICU care/death versus general ward admission. Multivariable models were developed using stepwise selection in logistic regression. *P*-values of <0.05 were considered statistically significant. We completed all analyses using SAS 9.4 (SAS Institute, Cary, NC).

## 3 RESULTS

Case data was collected for this study on 994 patients who presented to the ED and tested positive for COVID-19 between March 7 – August 9, 2020. Of these, 551 (55.4%) were discharged, 314 (31.6%) were admitted to the general ward, and 129 (13.0%) required ICU level care or died. Descriptive statistics for all three groups were compiled (Table 1). The most severely affected patient group had a significantly higher average age, had a higher percentage of males, and were more likely to be racial minorities. This group also had the highest percentage of patients with dyspnea, respiratory distress/failure and AMS. The group admitted to the general wards had the highest percentages of wet cough, chest pain, history of fever, fatigue/malaise, diarrhea, and syncope. The group of discharged patients had a higher percentage of patients with Hispanic or Latino ethnicity and had higher percentages of the symptoms of sore throat, dry cough, myalgia, headache, and olfactory/taste disturbance (Table 1).

**Table 1.** Characteristics of COVID-19 patients who presented at the GW Hospital Emergency Department in Washington, D.C.

| **Characteristic** | **Category 1**<br>**Discharge**<br>**(n=551)** | **Category 2**<br>**Hospital Admission**<br>**(n=314)** | **Category 3**<br>**ICU Care/Death**<br>**(n=129)** | **p-value\*** | **Total**<br>**(N=994)** |
| --- | --- | --- | --- | --- | --- |
| **Age, mean (SD)** | 43.7 (15.3) | 60.3 (15.6) | 68.4 (14.6) | <0.0001 | 52.1 (18.1) |
| **Age group, n (%)** |  |  |  |  |  |
| < 65 years | 502 (91.1) | 190 (60.7) | 45 (35.2) | <0.0001 | 737 (74.3) |
| ≥ 65 years | 49 (8.9) | 123 (39.3) | 83 (64.8) |  | 255 (25.7) |
| **Sex, n (%)** |  |  |  |  |  |
| Female | 281 (51.0) | 172 (54.8) | 52 (40.3) | 0.0215 | 505 (50.8) |
| Male | 270 (49.0) | 142 (45.2) | 77 (59.7) |  | 489 (49.2) |
| **Race, n (%)** |  |  |  |  |  |
| Black or African American | 306 (55.5) | 227 (72.3) | 94 (72.9) | <0.0001 | 627 (63.1) |
| White | 60 (10.9) | 16 (5.1) | 6 (4.7) |  | 82 (8.3) |
| Asian, American Indian, Alaska Native, Other | 185 (33.6) | 71 (22.6) | 29 (22.5) |  | 285 (28.7) |
| **Ethnicity, n (%)** |  |  |  |  |  |
| Non-Hispanic or -Latino | 412 (77.0) | 263 (85.9) | 108 (85.7) | 0.0022 | 783 (81.0) |
| Hispanic or Latino | 123 (23.0) | 43 (14.1) | 18 (14.3) |  | 184 (19.0) |
| **Sore throat, n (%)** |  |  |  |  |  |
| Yes | 82 (14.9) | 17 (5.4) | 5 (3.9) | <0.0001 | 104 (10.5) |
| No | 469 (85.1) | 297 (94.6) | 124 (96.1) |  | 890 (89.5) |
| **Cough without sputum (dry), n (%)** |  |  |  |  |  |
| Yes | 299 (54.3) | 153 (48.7) | 53 (41.1) | 0.0178 | 505 (50.8) |
| No | 252 (45.7) | 161 (51.3) | 76 (58.9) |  | 489 (49.2) |
| **Cough with sputum (wet), n (%)** |  |  |  |  |  |
| Yes | 47 (8.5) | 43 (13.7) | 9 (7.0) | 0.0245 | 99 (10.0) |
| No | 504 (91.5) | 271 (86.3) | 120 (93.0) |  | 895 (90.0) |
| **Chest pain, n (%)** |  |  |  |  |  |
| Yes | 125 (22.7) | 92 (29.3) | 16 (12.4) | 0.0006 | 233 (23.4) |
| No | 426 (77.3) | 222 (70.7) | 113 (87.6) |  | 761 (76.6) |
| **Shortness of breath/Dyspnea, n (%)** |  |  |  |  |  |
| Yes | 203 (36.8) | 187 (59.5) | 80 (62.0) | <0.0001 | 470 (47.3) |
| No | 348 (63.2) | 127 (40.5) | 49 (38.0) |  | 524 (52.7) |
| **Respiratory distress/failure, n (%)** |  |  |  |  |  |
| Yes | 3 (0.5) | 16 (5.1) | 20 (15.5) | <0.0001 | 39 (3.9) |
| No | 548 (99.5) | 298 (94.9) | 109 (84.5) |  | 955 (96.1) |
| **History of fever, n (%)** |  |  |  |  |  |
| Yes | 287 (52.1) | 166 (52.9) | 50 (38.8) | 0.0152 | 503 (50.6) |
| No | 264 (47.9) | 148 (47.1) | 79 (61.2) |  | 491 (49.4) |
| **Fatigue/Malaise, n (%)** |  |  |  |  |  |
| Yes | 170 (30.9) | 136 (43.3) | 36 (27.9) | 0.0003 | 342 (34.4) |
| No | 381 (69.1) | 178 (56.7) | 93 (72.1) |  | 652 (65.6) |
| **Muscle aches/Myalgia, n (%)** |  |  |  |  |  |
| Yes | 208 (37.8) | 66 (21.0) | 15 (11.6) | <0.0001 | 289 (29.1) |
| No | 343 (62.2) | 248 (79.0) | 114 (88.4) |  | 705 (70.9) |
| **Headache, n (%)** |  |  |  |  |  |
| Yes | 167 (30.3) | 48 (15.3) | 3 (2.3) | <0.0001 | 218 (21.9) |
| No | 384 (69.7) | 266 (84.7) | 126 (97.7) |  | 776 (78.1) |
| **Altered mental status/Confusion, n (%)** |  |  |  |  |  |
| Yes | 7 (1.3) | 45 (14.3) | 48 (37.2) | <0.0001 | 100 (10.1) |
| No | 544 (98.7) | 269 (85.7) | 81 (62.8) |  | 894 (89.9) |
| **Abdominal pain, n (%)** |  |  |  |  |  |
| Yes | 64 (11.6) | 38 (12.1) | 11 (8.5) | 0.5394 | 113 (11.4) |
| No | 487 (88.4) | 276 (87.9) | 118 (91.5) |  | 881 (88.6) |
| **Vomiting/Nausea, n (%)** |  |  |  |  |  |
| Yes | 111 (20.2) | 80 (25.5) | 25 (19.4) | 0.1477 | 216 (21.7) |
| No | 440 (79.8) | 234 (74.5) | 104 (80.6) |  | 778 (78.3) |
| **Diarrhea, n (%)** |  |  |  |  |  |
| Yes | 109 (19.8) | 92 (29.3) | 22 (17.0) | 0.0016 | 223 (22.4) |
| No | 442 (80.2) | 222 (70.7) | 107 (83.0) |  | 771 (77.6) |
| **Olfactory taste disturbance, n (%)** |  |  |  |  |  |
| Yes | 87 (15.8) | 19 (6.1) | 8 (6.2) | <0.0001 | 114 (11.5) |
| No | 464 (84.2) | 295 (93.9) | 121 (93.8) |  | 880 (88.5) |
| **Syncope, n (%)** |  |  |  |  |  |
| Yes | 10 (1.8) | 13 (4.1) | 5 (3.9) | 0.0887 | 28 (2.8) |
| No | 841 (98.2) | 301 (95.9) | 124 (96.1) |  | 966 (97.2) |

Abbreviations: COVID-19, coronavirus disease 2019; GW, The George Washington University; D.C., District of Columbia; ICU, intensive care unit; SD, standard deviation; ED, emergency department.
\* *P*-values were calculated using Chi-square/Fisher's Exact tests or ANOVA based on type of data.

When analyzed by multivariable logistic regression, the symptoms of dyspnea (aOR 2.7, 95% CI: 2.0-3.7) and diarrhea (aOR 1.6, 95% CI: 1.1-2.2) were independent predictors of hospital admission. Patient characteristics associated with hospital admission were age ≥ 65 years (aOR 7.4, 95% CI: 5.0-10.8) or being Black or African-American (aOR 3.0, 95% CI: 1.6-5.8) or Asian, American Indian, Alaska Native, or other race (aOR 2.2, 95% CI: 1.1-4.3). The symptoms of fatigue (OR 1.4, 95% CI: 1.1-1.9) and syncope (OR 2.3, 95% CI: 1.1-5.0) were associated with hospital admission by univariate analysis but were not independent predictors upon multivariable analysis (Table 2). The symptoms found to be negatively associated with admission and more likely to indicate discharge from the ED for convalescence at home were sore throat (aOR 0.4, 95% CI: 0.2-0.6), myalgia (aOR 0.5, 95% CI: 0.4-0.7), headache (aOR 0.5, 95% CI: 0.4-0.8), and olfactory/taste disturbance (aOR 0.5, 95% CI: 0.3-0.8). A Hispanic or Latino ethnicity was also negatively associated with hospital admission (OR 0.6, 95% CI: 0.4-0.8) but was not an independent predictor (Table 2).

**Table 2:**
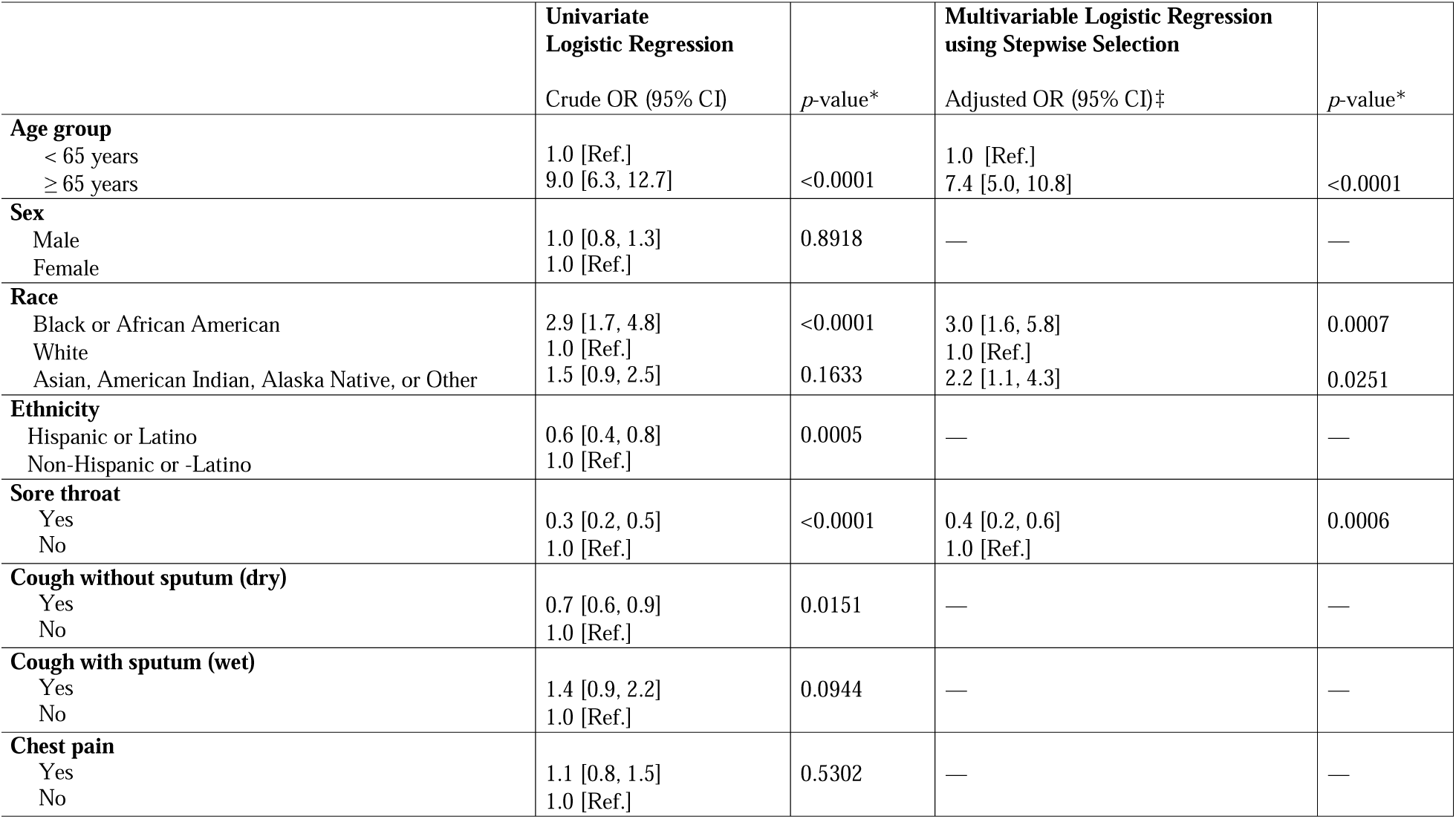
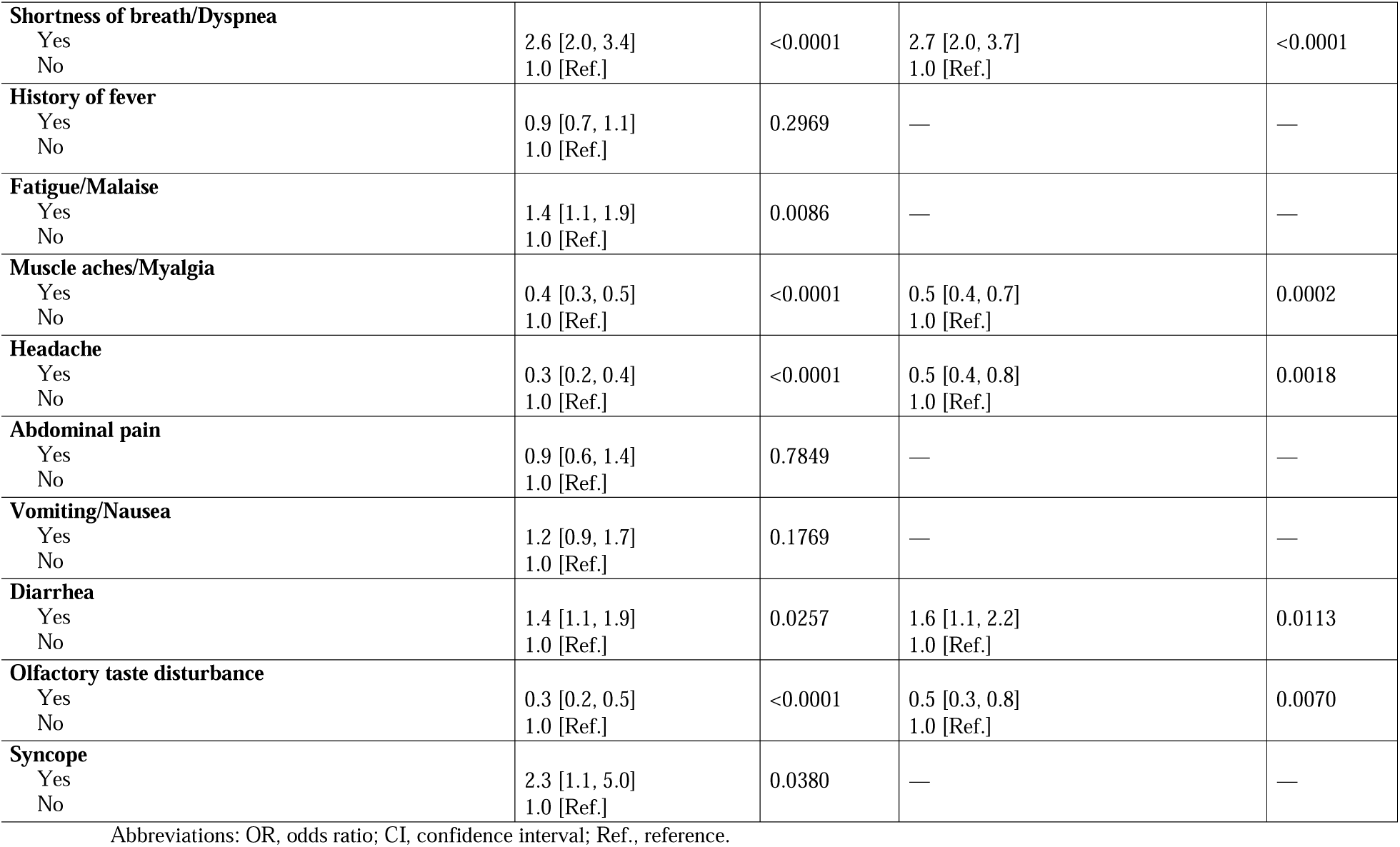
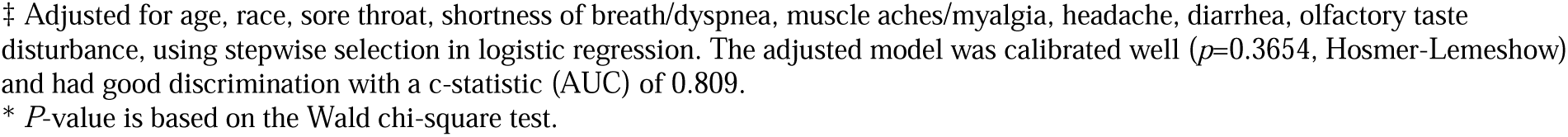
Logistic Regression Models for Hospital Admission/Death vs. Discharge

|  | Univariate<br>Logistic Regression |  | Multivariable Logistic Regression<br>using Stepwise Selection |  |
| --- | --- | --- | --- | --- |
|  | Crude OR (95% CI) | *p*-value* | Adjusted OR (95% CI)‡ | *p*-value* |
| **Age group** |  |  |  |  |
| < 65 years | 1.0 [Ref.] |  | 1.0 [Ref.] |  |
| ≥ 65 years | 9.0 [6.3, 12.7] | <0.0001 | 7.4 [5.0, 10.8] | <0.0001 |
| **Sex** |  |  |  |  |
| Male | 1.0 [0.8, 1.3] | 0.8918 | — | — |
| Female | 1.0 [Ref.] |  |  |  |
| **Race** |  |  |  |  |
| Black or African American | 2.9 [1.7, 4.8] | <0.0001 | 3.0 [1.6, 5.8] | 0.0007 |
| White | 1.0 [Ref.] |  | 1.0 [Ref.] |  |
| Asian, American Indian, Alaska Native, or Other | 1.5 [0.9, 2.5] | 0.1633 | 2.2 [1.1, 4.3] | 0.0251 |
| **Ethnicity** |  |  |  |  |
| Hispanic or Latino | 0.6 [0.4, 0.8] | 0.0005 | — | — |
| Non-Hispanic or -Latino | 1.0 [Ref.] |  |  |  |
| **Sore throat** |  |  |  |  |
| Yes | 0.3 [0.2, 0.5] | <0.0001 | 0.4 [0.2, 0.6] | 0.0006 |
| No | 1.0 [Ref.] |  | 1.0 [Ref.] |  |
| **Cough without sputum (dry)** |  |  |  |  |
| Yes | 0.7 [0.6, 0.9] | 0.0151 | — | — |
| No | 1.0 [Ref.] |  |  |  |
| **Cough with sputum (wet)** |  |  |  |  |
| Yes | 1.4 [0.9, 2.2] | 0.0944 | — | — |
| No | 1.0 [Ref.] |  |  |  |
| **Chest pain** |  |  |  |  |
| Yes | 1.1 [0.8, 1.5] | 0.5302 | — | — |
| No | 1.0 [Ref.] |  |  |  |
| **Shortness of breath/Dyspnea**<br>Yes<br>No | 2.6 [2.0, 3.4]<br>1.0 [Ref.] | <0.0001 | 2.7 [2.0, 3.7]<br>1.0 [Ref.] | <0.0001 |
| **History of fever**<br>Yes<br>No | 0.9 [0.7, 1.1]<br>1.0 [Ref.] | 0.2969 | — | — |
| **Fatigue/Malaise**<br>Yes<br>No | 1.4 [1.1, 1.9]<br>1.0 [Ref.] | 0.0086 | — | — |
| **Muscle aches/Myalgia**<br>Yes<br>No | 0.4 [0.3, 0.5]<br>1.0 [Ref.] | <0.0001 | 0.5 [0.4, 0.7]<br>1.0 [Ref.] | 0.0002 |
| **Headache**<br>Yes<br>No | 0.3 [0.2, 0.4]<br>1.0 [Ref.] | <0.0001 | 0.5 [0.4, 0.8]<br>1.0 [Ref.] | 0.0018 |
| **Abdominal pain**<br>Yes<br>No | 0.9 [0.6, 1.4]<br>1.0 [Ref.] | 0.7849 | — | — |
| **Vomiting/Nausea**<br>Yes<br>No | 1.2 [0.9, 1.7]<br>1.0 [Ref.] | 0.1769 | — | — |
| **Diarrhea**<br>Yes<br>No | 1.4 [1.1, 1.9]<br>1.0 [Ref.] | 0.0257 | 1.6 [1.1, 2.2]<br>1.0 [Ref.] | 0.0113 |
| **Olfactory taste disturbance**<br>Yes<br>No | 0.3 [0.2, 0.5]<br>1.0 [Ref.] | <0.0001 | 0.5 [0.3, 0.8]<br>1.0 [Ref.] | 0.0070 |
| **Syncope**<br>Yes<br>No | 2.3 [1.1, 5.0]<br>1.0 [Ref.] | 0.0380 | — | — |

Abbreviations: OR, odds ratio; CI, confidence interval; Ref., reference.
‡ Adjusted for age, race, sore throat, shortness of breath/dyspnea, muscle aches/myalgia, headache, diarrhea, olfactory taste disturbance, using stepwise selection in logistic regression. The adjusted model was calibrated well ( $p=0.3654$ , Hosmer-Lemeshow) and had good discrimination with a c-statistic (AUC) of 0.809.
\* $P$ -value is based on the Wald chi-square test.

The presenting symptom in the ED of AMS (aOR 3.8, 95% CI: 2.1-6.6) was the only symptom ultimately independently predictive with the development of severe COVID-19 requiring ICU care or resulting in eventual death as opposed to admission to the general wards. Respiratory distress (OR 3.4, 95% CI: 1.7-6.8), an age ≥ 65 years (OR 2.8. 95% CI: 1.9-4.4) and male sex (OR 1.8, 95% CI: 1.2-2.7) were associated with requiring ICU care but were not independently predictive (Table 3). A history of fever was the only symptom negatively predictive of patients requiring ICU care or dying and was more likely associated with remaining on the general wards (aOR 0.5, 95% CI: 0.3-0.8). Chest pain (OR 0.3, 95% CI: 0.2-0.6), fatigue (OR 0.5, 95% CI: 0.3-0.8), myalgia (OR 0.5, 95% CI: 0.3-0.9), and diarrhea (OR 0.5, 95% CI: 0.3-0.8) were also associated with only requiring admission to the general ward but were also not independent predictors (Table 3).

**Table 3.** Logistic Regression Models for ICU care/Death vs. General Ward Admission

|  | Univariate<br>Logistic Regression |  | Multivariable Logistic Regression<br>using Stepwise Selection |  |
| --- | --- | --- | --- | --- |
|  | Crude OR (95% CI) | *p*-value* | Adjusted OR (95% CI)‡ | *p*-value* |
| **Age group** |  |  |  |  |
| < 65 years | 1.0 [Ref.] |  | — | — |
| ≥ 65 years | 2.8 [1.9, 4.4] | <0.0001 |  |  |
| **Sex** |  |  |  |  |
| Male | 1.8 [1.2, 2.7] | 0.0059 | — | — |
| Female | 1.0 [Ref.] |  |  |  |
| **Race** |  |  |  |  |
| Black or African American | 1.1 [0.4, 2.9] | 0.8409 | — | — |
| White | 1.0 [Ref.] |  |  |  |
| Asian, American Indian, Alaska Native, or Other | 1.1 [0.4, 3.1] | 0.8712 |  |  |
| **Ethnicity** |  |  |  |  |
| Hispanic or Latino | 1.0 [0.6, 1.8] | 0.9494 | — | — |
| Non-Hispanic or -Latino | 1.0 [Ref.] |  |  |  |
| **Sore throat** |  |  |  |  |
| Yes | 0.7 [0.3, 2.0] | 0.5006 | — | — |
| No | 1.0 [Ref.] |  |  |  |
| **Cough without sputum (dry)** |  |  |  |  |
| Yes | 0.7 [0.5, 1.1] | 0.1436 | — | — |
| No | 1.0 [Ref.] |  |  |  |
| **Cough with sputum (wet)** |  |  |  |  |
| Yes | 0.5 [0.2, 1.0] | 0.0502 | — | — |
| No | 1.0 [Ref.] |  |  |  |
| **Chest pain** |  |  |  |  |
| Yes | 0.3 [0.2, 0.6] | 0.0003 | — | — |
| No | 1.0 [Ref.] |  |  |  |
| **Shortness of breath/Dyspnea** |  |  |  |  |
| Yes | 1.1 [0.7, 1.7] | 0.6306 | — | — |
| No | 1.0 [Ref.] |  |  |  |
| **Respiratory distress/failure** |  |  |  |  |
| Yes | 3.4 [1.7, 6.8] | 0.0005 | — | — |
| No | 1.0 [Ref.] |  |  |  |
| **History of fever** |  |  |  |  |
| Yes | 0.6 [0.4, 0.9] | 0.0073 | 0.5 [0.3, 0.8] | 0.0067 |
| No | 1.0 [Ref.] |  | 1.0 [Ref.] |  |
| **Fatigue/Malaise** |  |  |  |  |
| Yes | 0.5 [0.3, 0.8] | 0.0027 | — | — |
| No | 1.0 [Ref.] |  |  |  |
| **Muscle aches/Myalgia** |  |  |  |  |
| Yes | 0.5 [0.3, 0.9] | 0.0220 | — | — |
| No | 1.0 [Ref.] |  |  |  |
| **Altered mental status/Confusion** |  |  |  |  |
| Yes | 3.5 [2.2, 5.7] | <0.0001 | 3.8 [2.1, 6.6] | <0.0001 |
| No | 1.0 [Ref.] |  | 1.0 [Ref.] |  |
| **Abdominal pain** |  |  |  |  |
| Yes | 0.7 [0.3, 1.4] | 0.2784 | — | — |
| No | 1.0 [Ref.] |  |  |  |
| **Vomiting/Nausea** |  |  |  |  |
| Yes | 0.7 [0.4, 1.2] | 0.1717 | — | — |
| No | 1.0 [Ref.] |  |  |  |
| **Diarrhea** |  |  |  |  |
| Yes | 0.5 [0.3, 0.8] | 0.0081 | — | — |
| No | 1.0 [Ref.] |  |  |  |
| **Olfactory taste disturbance** |  |  |  |  |
| Yes | 1.0 [0.4, 2.4] | 0.9518 | — | — |
| No | 1.0 [Ref.] |  |  |  |
| **Syncope** |  |  |  |  |
| Yes | 0.9 [0.3, 2.7] | 0.8982 | — | — |
| No | 1.0 [Ref.] |  |  |  |

Abbreviations: ICU, intensive care unit; OR, odds ratio; CI, confidence interval; Ref., reference.
‡ Adjusted for history of fever and altered mental status/confusion, using stepwise selection in logistic regression. The adjusted model was calibrated well ( $p=0.6742$ , Hosmer-Lemeshow) and had fair discrimination with a c-statistic (AUC) of 0.696.
\* $P$ -value is based on the Wald chi-square test.

## 4 DISCUSSION

The clinical symptoms at ED presentation are associated with different degrees of clinical severity. The objective of this study was to examine the associations between presenting symptoms exhibited by symptomatic COVID-19 positive and the clinical outcomes for those patients. Respiratory failure and AMS were associated with the most severe clinical course, with AMS being the only independently predictive symptom. Chest pain, history of fever, fatigue, myalgia, and diarrhea were associated with moderate severity. Finally, sore throat, headache, myalgia, or an olfactory or taste disturbance were associated with the most benign severity as these patients were more likely to convalesce at home.

The comorbidities such as obesity, increased age, diabetes, and cardiovascular disease were associated with worse outcomes in COVID-19 patients and has been well-documented in prior literature. [4; 7-12] Fewer studies have established the associations between presenting symptomatology and subsequent patient prognoses. Our finding of AMS being an independent predictor of increased likelihood of ICU care or death is replicated in the literature. Analysis of a cohort of 710 hospitalized patients found AMS was independently associated with ICU admission and in-hospital mortality. [13] A study of COVID patients presenting to the ED with neurological symptoms found that altered mental status on admission was associated with increased mortality. [14] The association of respiratory symptoms with worse prognoses has also reported by several studies. A similar cohort of the first 1000 cases presenting to the ED of two hospitals in New York City found dyspnea was the only presenting symptom associated with patients eventually requiring ICU care, and nausea or vomiting were associated with less severe outcomes. [4] A cohort of 43,103 COVID-19 outpatients in France found that dyspnea was associated with an eventual requirement for hospitalization while anosmia or ageusia were associated with a decreased risk for need for hospitalization.[15] The same dyspnea and anosmia/ageusia results were found in a smaller cohort of initial outpatients in Seattle, Washington as well. [16] A meta-analysis encompassing 1813 patients revealed that dyspnea was associated with an increased risk for ICU admission (pooled OR 6.55, 95% CI: 4.28-10.0). [17] A retrospective study of 463 patients presenting to an ED in Detroit, Michigan reported that dyspnea, tachypnea, and hypoxia were the only symptoms associated with ICU requirement whereas fever and headache were more common among general ward patients than ICU patients. [10]

When considering whether a patient is a candidate to recover at home, the safety of discharging certain low-risk patients from the ED has been described previously. A prospective study of 452 discharged patients deemed low-risk demonstrated a re-admission rate of 4.6% and ICU admission rate of only 0.7%. In this study, low risk was defined as oxygen saturation ≥ 92%, respiratory rate ≤ 20 breaths/minute, ambulatory, age ≤ 60 years, the lack of significant comorbidities, mild or no disease on chest radiograph, and the ability to return within 24 hours. [18] A retrospective study of 199 COVID-19 positive patients discharged from an ED in London, England found a 6% rate of subsequent hospitalization. [19] A prospective study of 314 low-risk COVID-19 patients who had an eventual admission rate of only 6.4%.[20] Low rates of readmissions demonstrate that ED physicians are successfully risk-stratifying COVID-19 patients even with limited information.

### 4.1 Strengths/Limitations

Strengths of our study include a high quality chart review for all of the patients included in the study, along with a 30 day buffer period after the patients’ index ED presentation to check for any subsequent admissions, re-admissions, or worsening clinical course. We reclassified patients into different severity categories based on worsening disease course and the requirement of elevated levels of care. This helps to account for the unpredictable clinical course that COVID-19 may assume in different patients. Furthermore, the three categories of severity are pragmatic as they are the three most common dispositions for patients after ED evaluation and as such, are based on the needs of the patient.

Although the three categories are delineated along the three strata of patient care, one limitation of the study is that the categories of severity were still dependent on individual physician decision regarding the necessary level of care. Due to the fluctuating prevalence of COVID-19 patients over the course of the pandemic, this decision may also have been influenced by hospital system factors such as crowding or surges in the patient census. Furthermore, due to the evolving clinical care and guidance over the year of the pandemic, there may have been shifting standards for admission. Our study was a single-center retrospective study, so there may be variation in generalization to other institutions due to different patient populations or institutional care guidelines.

Several limitations should be highlighted for this study. First, although each patient received rigorous and standardized chart review, certain minor symptoms may have been under-reported due to clinical severity (e.g., anosmia may not have been reported if the patient is also in respiratory distress). Second, due to the recent rapid onset of the pandemic, extended long-term outcomes were not able to be included in the definition of severe versus non-severe outcomes. Only the level of in-hospital care or discharge were used as categorical outcomes for analysis. In addition, relative disability or quality of life after discharge were not factored into the outcome analysis. Similarly, we were unable to account for any patients who were discharged and but then were admitted to a different hospital or died at home. Finally, we also were unable to measure relative patient success of different measures to assist convalescence at home. For example, whether or not patients with home pulse-ox measuring or access to telemedicine had better outcomes than those without.

Our cohort of patients in this study includes only those who presented to the ED with symptomatic COVID-19. Patients exhibiting milder symptoms may have opted to initially present to primary care or urgent care settings for testing rather than the ED, which could lead to an overall cohort of comparatively sicker patients in our study as opposed to the general population of COVID-19 positive patients. Additionally, due to the standardized nature of the data collection protocols as our study was part of a larger multi-center study, we were not able to quantify the relative severity of patient comorbidities and how those might impact the presentation of symptoms in individual patients. Lastly, due to lack of standardized treatment guidelines for COVID-19, especially in the early stages of the pandemic, in-hospital treatment for patients was not able to be controlled for across the cohort.

Our study demonstrates associations between symptoms of COVID patients upon presentation in the ED and clinical outcomes. Further research is needed to understand the time-course for disease progression and to develop integrated prediction scores for 30-day outcomes.

## 5 CONCLUSIONS

This study shows that initial presenting symptoms of respiratory failure and AMS in ED patients with COVID-19 were associated with a severe disease course. Symptoms of a history of fever, chest pain, myalgia, fatigue, and diarrhea were associated with a moderate disease course. Anosmia, ageusia, sore throat, headache, and myalgia were associated with the most benign disease course as patients were more likely to successfully convalesce at home. COVID-19 presents with a heterogeneous constellation of symptoms, and a better understanding of the association of the presenting symptoms within the ED with the ultimate patient prognoses may be useful for allocating resources and targeting management plans.

## Data Availability

Data is currently at the lead site, Indiana University

## REFERENCES

[1] COVID data tracker. Centers for Disease Control and Prevention Web site. https://covid.cdc.gov/covid-data-tracker. Updated 2020. Accessed June 3, 2021.

[2] Goyal P, Choi JJ, Pinheiro LC, et al. Clinical characteristics of covid-19 in new york city. N Engl J Med. 2020;382(24):2372–2374. https://doi.org/10.1056/NEJMc2010419. doi: 10.1056/NEJMc2010419.

[3] Docherty AB, Harrison EM, Green CA, et al. Features of 20□133 UK patients in hospital with covid-19 using the ISARIC WHO clinical characterisation protocol: Prospective observational cohort study. BMJ. 2020;369. https://www.ncbi.nlm.nih.gov/pmc/articles/PMC7243036/. Accessed Sep 16, 2020. doi: 10.1136/bmj.m1985.

[4] Argenziano MG, Bruce SL, Slater CL, et al. Characterization and clinical course of 1000 patients with coronavirus disease 2019 in new york: Retrospective case series. BMJ. 2020;369:m1996. Accessed Sep 16, 2020. doi: 10.1136/bmj.m1996.

[5] Li L, Huang T, Wang Y, et al. COVID-19 patients’ clinical characteristics, discharge rate, and fatality rate of meta-analysis. J Med Virol. 2020;92(6):577–583. Accessed Sep 15, 2020. doi: 10.1002/jmv.25757.

[6] Gilbert EH, Lowenstein SR, Koziol-McLain J, Barta DC, Steiner J. Chart reviews in emergency medicine research: Where are the methods? Ann Emerg Med. 1996;27(3):305–308. Accessed Oct 29, 2020. doi: 10.1016/s0196-0644(96)70264-0.

[7] Palaiodimos L, Kokkinidis DG, Li W, et al. Severe obesity, increasing age and male sex are independently associated with worse in-hospital outcomes, and higher in-hospital mortality, in a cohort of patients with COVID-19 in the bronx, new york. Metab Clin Exp. 2020;108:154262. Accessed Sep 16, 2020. doi: 10.1016/j.metabol.2020.154262.

[8] Aggarwal G, Cheruiyot I, Aggarwal S, et al. Association of cardiovascular disease with coronavirus disease 2019 (COVID-19) severity: A meta-analysis. Curr Probl Cardiol. 2020;45(8):100617. Accessed Sep 16, 2020. doi: 10.1016/j.cpcardiol.2020.100617.

[9] Targher G, Mantovani A, Wang X-, et al. Patients with diabetes are at higher risk for severe illness from COVID-19. Diabetes Metab. 2020;46(4):335–337. https://www.ncbi.nlm.nih.gov/pmc/articles/PMC7255326/. Accessed Oct 3, 2020. doi: 10.1016/j.diabet.2020.05.001.

[10] Suleyman G, Fadel RA, Malette KM, et al. Clinical characteristics and morbidity associated with coronavirus disease 2019 in a series of patients in metropolitan detroit. JAMA Netw Open. 2020;3(6). https://www.ncbi.nlm.nih.gov/pmc/articles/PMC7298606/. Accessed Sep 18, 2020. doi: 10.1001/jamanetworkopen.2020.12270.

[11] Ebinger JE, Achamallah N, Ji H, et al. Pre-existing traits associated with covid-19 illness severity. PLoS One. 2020;15(7). https://www.ncbi.nlm.nih.gov/pmc/articles/PMC7377468/. Accessed Oct 3, 2020. doi: 10.1371/journal.pone.0236240.

[12] Bennett KE, Mullooly M, O’Loughlin M, et al. Underlying conditions and risk of hospitalisation, ICU admission and mortality among those with COVID-19 in ireland: A national surveillance study. Lancet Reg Health Eur. 2021;5:100097. Accessed May 5, 2021. doi: 10.1016/j.lanepe.2021.100097.

[13] Kenerly MJ, Shah P, Patel H, et al. Altered mental status is an independent predictor of mortality in hospitalized COVID-19 patients. Ir J Med Sci. 2021. Accessed Jun 3, 2021. doi: 10.1007/s11845-021-02515-4.

[14] García-Azorín D, Trigo J, Martínez-Pías E, et al. Neurological symptoms in covid-19 patients in the emergency department. Brain Behav. 2021;11(4):e02058. Accessed Jun 3, 2021. doi: 10.1002/brb3.2058.

[15] Yordanov Y, Dinh A, Bleibtreu A, et al. Clinical characteristics and factors associated with hospital admission or death in 43,103 adult outpatients with COVID-19 managed with the covidom telesurveillance solution: A prospective cohort study. Clinical microbiology and infection. 2021;0(0). https://www.ncbi.nlm.nih.gov/pubmed/33915287. doi: 10.1016/j.cmi.2021.04.010.

[16] Ylescupidez A, Rips A, Bahnson HT, et al. Early prognostic indicators of subsequent hospitalization in patients with mild COVID-19. J Clin Med. 2021;10(8). https://www.ncbi.nlm.nih.gov/pmc/articles/PMC8068070/. Accessed May 5, 2021. doi: 10.3390/jcm10081562.

[17] Jain V, Yuan J. Predictive symptoms and comorbidities for severe COVID-19 and intensive care unit admission: A systematic review and meta-analysis. Int J Public Health. 2020;65(5):533–546. Accessed Sep 15, 2020. doi: 10.1007/s00038-020-01390-7.

[18] Berdahl CT, Glennon NC, Henreid AJ, Torbati SS. The safety of home discharge for low□risk emergency department patients presenting with coronavirus□like symptoms during the COVID□19 pandemic: A retrospective cohort study. J Am Coll Emerg Physicians Open. 2020. https://www.ncbi.nlm.nih.gov/pmc/articles/PMC7436406/. Accessed Sep 15, 2020. doi: 10.1002/emp2.12230.

[19] Lanham D, Roe J, Chauhan A, et al. COVID-19 emergency department discharges: An outcome study. Clinical medicine (London, England). 2021;21(2):e126–e131. https://www.ncbi.nlm.nih.gov/pubmed/33419864. doi: 10.7861/clinmed.2020-0817.

[20] Francisco Javier Teigell Muñoz, · Elena García-Guijarro, · Paula García-Domingo, et al. A safe protocol to identify low-risk patients with COVID-19 pneumonia for outpatient management. Intern Emerg Med. 2021:1–9. https://www.ncbi.nlm.nih.gov/pmc/articles/PMC7900647/. Accessed May 5, 2021. doi: 10.1007/s11739-021-02660-9.

